# Diagnostic utility of a Ferritin-to-Procalcitonin Ratio to differentiate patients with COVID-19 from those with Bacterial Pneumonia: A multicenter study

**DOI:** 10.1101/2020.10.20.20216309

**Authors:** Amal A. Gharamti, Fei Mei, Katherine C. Jankousky, Jin Huang, Peter Hyson, Daniel B. Chastain, Jiawei Fan, Sharmon Osae, Wayne W. Zhang, José G. Montoya, Kristine M. Erlandson, Sias J. Scherger, Carlos Franco-Paredes, Andrés F. Henao-Martínez, Leland Shapiro

## Abstract

**Importance:** There is a need to develop tools to differentiate COVID-19 from bacterial pneumonia at the time of clinical presentation before diagnostic testing is available.

**Objective:** To determine if the Ferritin-to-Procalcitonin ratio (F/P) can be used to differentiate COVID-19 from bacterial pneumonia.

**Design:** This case-control study compared patients with either COVID-19 or bacterial pneumonia, admitted between March 1 and May 31, 2020. Patients with COVID-19 and bacterial pneumonia co-infection were excluded.

**Setting:** A multicenter study conducted at three hospitals that included UCHealth and Phoebe Putney Memorial Hospital in the United States, and Yichang Central People’s Hospital in China.

**Participants:** A total of 242 cases with COVID-19 infection and 34 controls with bacterial pneumonia.

**Main Outcomes and Measures:** The F/P in patients with COVID-19 or with bacterial pneumonia were compared. Receiver operating characteristic analysis determined the sensitivity and specificity of various cut-off F/P values for the diagnosis of COVID-19 versus bacterial pneumonia.

**Results:** Patients with COVID-19 pneumonia had a lower mean age (57.11 vs 64.4 years, p=0.02) and a higher BMI (30.74 vs 27.15 kg/m^2^, p=0.02) compared to patients with bacterial pneumonia. Cases and controls had a similar proportion of women (47% vs 53%, p=0.5) and COVID-19 patients had a higher prevalence of diabetes mellitus (32.6% vs 12%, p=0.01). The median F/P was significantly higher in patients with COVID-19 (4037.5) compared to the F/P in bacterial pneumonia (802, p<0.001). An F/P ≥ 877 used to diagnose COVID-19 resulted in a sensitivity of 85% and a specificity of 56%, with a positive predictive value of 93.2%, and a likelihood ratio of 1.92. In multivariable analyses, an F/P ≥ 877 was associated with greater odds of identifying a COVID-19 case (OR: 11.27, CI: 4-31.2, p<0.001).

**Conclusions and Relevance:** An F/P ≥ 877 increases the likelihood of COVID-19 pneumonia compared to bacterial pneumonia. Further research is needed to determine if obtaining ferritin and procalcitonin simultaneously at the time of clinical presentation has improved diagnostic value. Additional questions include whether an increased F/P and/or serial F/P associates with COVID-19 disease severity or outcomes.

## Introduction

The Coronavirus Disease 2019 (COVID-19) was declared a global pandemic by the World Health Organization on March 11, 2020.^1^ It is challenging to differentiate COVID-19 pneumonia from bacterial pneumonia at the time of clinical presentation. This difficulty is underscored in clinical series of COVID-19 patients that document concomitant use of antibiotics in up to 100% of COVID-19 patients.^2-4^ In 4,267 COVID-19 patients in New York City, co-infections were diagnosed in 3.6% of the patients (mostly bacterial), yet antibiotics were given in 71% of these patients.^5^ This mismatch between true co-infection and clinically-presumed co-infection emphasizes the difficulty in separating COVID-19 from bacterial infection. Early diagnosis enables expeditious use of appropriate antiviral or antibacterial treatments, minimizes adverse effects and costs associated with empirical therapies targeting both viral and bacterial infection, and facilitates infection control measures designed to prevent COVID-19 transmission. Real-Time Reverse Transcriptase Polymerase Chain Reaction (RT-PCR) used to diagnose COVID-19 typically requires a minimum 24 hours to return and can take more than 72 hours in low-resource settings. The clinical utility of other rapid antigen or molecular-based tests has not been established.^6^ Thus, investigation of biomarkers has emerged as an important area of study in efforts to discriminate COVID-19 pneumonia from bacterial pneumonia.

Several laboratory markers have been studied for use in diagnosis and assessing severity of COVID-19 at the time of clinical presentation. We employed a model-based approach that led us to focus on procalcitonin (PCT) and the iron-storage protein ferritin. Our model predicts the ratio of circulating ferritin level (ng/ml) and PCT (ng/ml), called the ferritin/procalcitonin ratio (F/P), would be higher in COVID-19 pneumonia than in bacterial pneumonia.

## Methods

### Ethics statement

This investigation was approved by the Colorado Multiple Institutional Review Board (COMIRB) at the University of Colorado Anschutz Medical Campus (UCHealth), the Institutional Review Board (IRB) at Phoebe Putney Memorial Hospital in Albany, Georgia (USA), and the IRB at Yichang Central People’s Hospital of China Three Gorges University. Clinical data were obtained under a protocol reviewed and approved by IRBs and ethics committees at UCHealth (COMIRB Protocol 20-0690), Phoebe Putney Memorial Hospital (PROJECT00002679), and Yichang Central People’s Hospital (HEC-KYJJ-2020-026-01).

### Patients and data collection

COVID-19 patients admitted to UCHealth, Yichang Central People’s Hospital, or Phoebe Putney Memorial Hospital from March 1 to May 31, 2020, were considered for inclusion. The diagnosis of COVID-19 pneumonia was established by positive RT-PCR along with signs, symptoms, and imaging findings suggesting pneumonia. Exclusion criteria included patients younger than 18 years, pregnancy, death on admission, missing baseline data, or being transferred to other hospitals. We excluded patients with concurrent COVID-19 and bacterial pneumonia. Patients diagnosed with bacterial pneumonia in the absence of COVID-19 served as controls. Bacterial pneumonia was diagnosed by clinical signs and symptoms of pneumonia, radiologic evidence of pneumonia on chest imaging, microbiologic documentation of a bacterial pathogen, and a negative COVID-19 PCR test.

Electronic medical records were manually abstracted for clinical and laboratory variables. At UCHealth and Phoebe Putney Memorial Hospital, data were retrospectively collected and stored in RedCap (electronic capture tools). At Yichang Central People’s Hospital, data were transferred manually into an excel file. The following data were collected: gender, race/ethnicity, age, weight, and body mass index (BMI), oxygen saturation in the Emergency Department, diagnosed pre-existing medical conditions including diabetes mellitus (DM) and hypertension (HTN), and laboratory results including lactate dehydrogenase (LDH), ferritin, and PCT.

### Real-Time Reverse Transcriptase Polymerase Chain Reaction (RT-PCR)

At UCHealth, upper respiratory samples were subjected to reverse transcription PCR amplification for Severe Acute Respiratory Syndrome Coronavirus 2 (SARS-CoV-2, Roche Cobas 6800 EUA, Roche Diagnostics Corporation, Indianapolis, Indiana).

At Phoebe Putney Memorial Hospital, several PCR platforms were used, including tests employed at Labcorp, Quest, and use of the Cepheid Xpert® Xpress SARS-CoV-2 (Cepheid, Sunnyvale, California, USA).

At Yichang Central People’s Hospital, RT-PCR was performed using the Detection Kit for SARS-CoV-2 RNA (PCR-Fluorescence Probing, Da An Gene Co., Ltd. of Sun Yat-sen University, Guandong, China) to detect SARS-CoV-2 virus nucleic acid.

### Statistical analysis

Statistical analyses were performed using STATA software, version 12.1 (StataCorp, College Station, Texas, USA). Continuous variables were summarized using means (± standard deviations) or medians (± interquartile ranges), and frequencies (percent) were used for categorical variables. Dichotomous outcomes were compared using chi-square. Fisher exact test was used for nominal independent variables, and t-test or Mann-Whitney U test were employed for interval independent variables. Co-linear variables or variables with missing significant data were excluded. We excluded from analysis 102 cases with COVID-19 and bacterial pneumonia co-infections. We compared F/P in patients diagnosed with COVID-19 to F/P in patients with bacterial pneumonia using a logistic regression controlling for age, sex, BMI, DM, HTN, and cohort site. We interrogated the receiver operating characteristic (ROC) curve that compared true positive rates (sensitivity) to false-positive rates (1- specificity) for various F/P values. We determined the optimal F/P value that separated patients with COVID-19 pneumonia from patients with bacterial pneumonia. Statistical significance was set at the 0.05 alpha level.

We analyzed F/P in the UCHealth cohort and separately analyzed F/P in the three hospitals combined. A ROC curve was constructed for F/P in UCHealth patients and separately in the three hospital cohorts combined, and areas under the ROC curves calculated.

### Data access

The corresponding author had full access to data in the study and had final responsibility for the decision to submit the manuscript for publication. The datasets used in the current study are available from the corresponding author on a reasonable request.

## Results

### Clinical characteristics and laboratory data in the three participating sites

Table 1 shows the clinical characteristics and laboratory data obtained at each participating hospital. DM and HTN were most prevalent in the Phoebe Putney Memorial Hospital group. A significantly larger percentage of patients from UCHealth and Phoebe Putney Memorial Hospitals were intubated compared to patients from the Yichang Central People’s Hospital. The median ferritin and procalcitonin levels were higher in Phoebe Putney Memorial Hospital’s patients compared to patients at UCHealth and Yichang Central People’s Hospital. The median F/P was lower in the UCHealth cohort compared to the cohorts from Phoebe Putney Memorial Hospital and Yichang Central People’s Hospital.

**Table 1:**
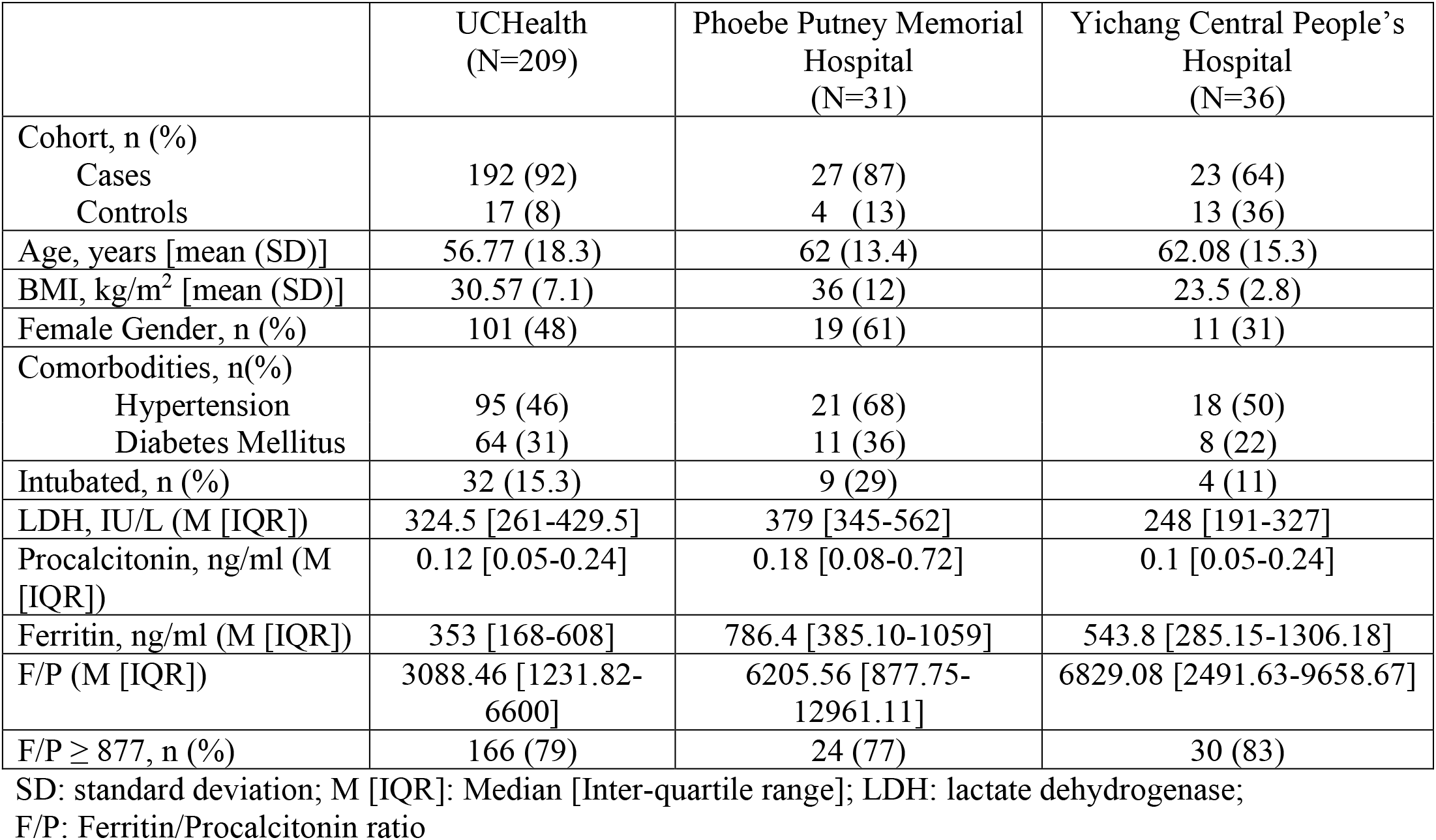
Characteristics of patients included at the three participating sites

|  | UCHealth<br>(N=209) | Phoebe Putney Memorial<br>Hospital<br>(N=31) | Yichang Central People's<br>Hospital<br>(N=36) |
| --- | --- | --- | --- |
| Cohort, n (%) |  |  |  |
| Cases | 192 (92) | 27 (87) | 23 (64) |
| Controls | 17 (8) | 4 (13) | 13 (36) |
| Age, years [mean (SD)] | 56.77 (18.3) | 62 (13.4) | 62.08 (15.3) |
| BMI, kg/m<sup>2</sup> [mean (SD)] | 30.57 (7.1) | 36 (12) | 23.5 (2.8) |
| Female Gender, n (%) | 101 (48) | 19 (61) | 11 (31) |
| Comorbidities, n(%) |  |  |  |
| Hypertension | 95 (46) | 21 (68) | 18 (50) |
| Diabetes Mellitus | 64 (31) | 11 (36) | 8 (22) |
| Intubated, n (%) | 32 (15.3) | 9 (29) | 4 (11) |
| LDH, IU/L (M [IQR]) | 324.5 [261-429.5] | 379 [345-562] | 248 [191-327] |
| Procalcitonin, ng/ml (M [IQR]) | 0.12 [0.05-0.24] | 0.18 [0.08-0.72] | 0.1 [0.05-0.24] |
| Ferritin, ng/ml (M [IQR]) | 353 [168-608] | 786.4 [385.10-1059] | 543.8 [285.15-1306.18] |
| F/P (M [IQR]) | 3088.46 [1231.82-6600] | 6205.56 [877.75-12961.11] | 6829.08 [2491.63-9658.67] |
| F/P ≥ 877, n (%) | 166 (79) | 24 (77) | 30 (83) |

SD: standard deviation; M [IQR]: Median [Inter-quartile range]; LDH: lactate dehydrogenase;
F/P: Ferritin/Procalcitonin ratio

### Characteristics of patients with COVID-19 and patients with bacterial pneumonia

We evaluated 242 hospitalized patients diagnosed with COVID-19 and 34 hospitalized patients diagnosed with bacterial pneumonia in the three participating hospitals combined, and patient characteristics are shown in Table 2 (none of these patients were co-infected). Patients with COVID-19 pneumonia were significantly younger and had a significantly higher mean BMI than patients with bacterial pneumonia. Both groups had similar proportions of women. DM was significantly more common in the COVID-19 group compared to the group with bacterial pneumonia. Mean oxygen saturation in COVID-19 patients upon presentation in the Emergency Department was lower than that in patients with bacterial pneumonia, although the difference was not statistically significant. Patients with COVID-19 pneumonia were more likely to have a course complicated by intubation compared to patients with bacterial pneumonia, without reaching statistical significance.

**Table 2:**
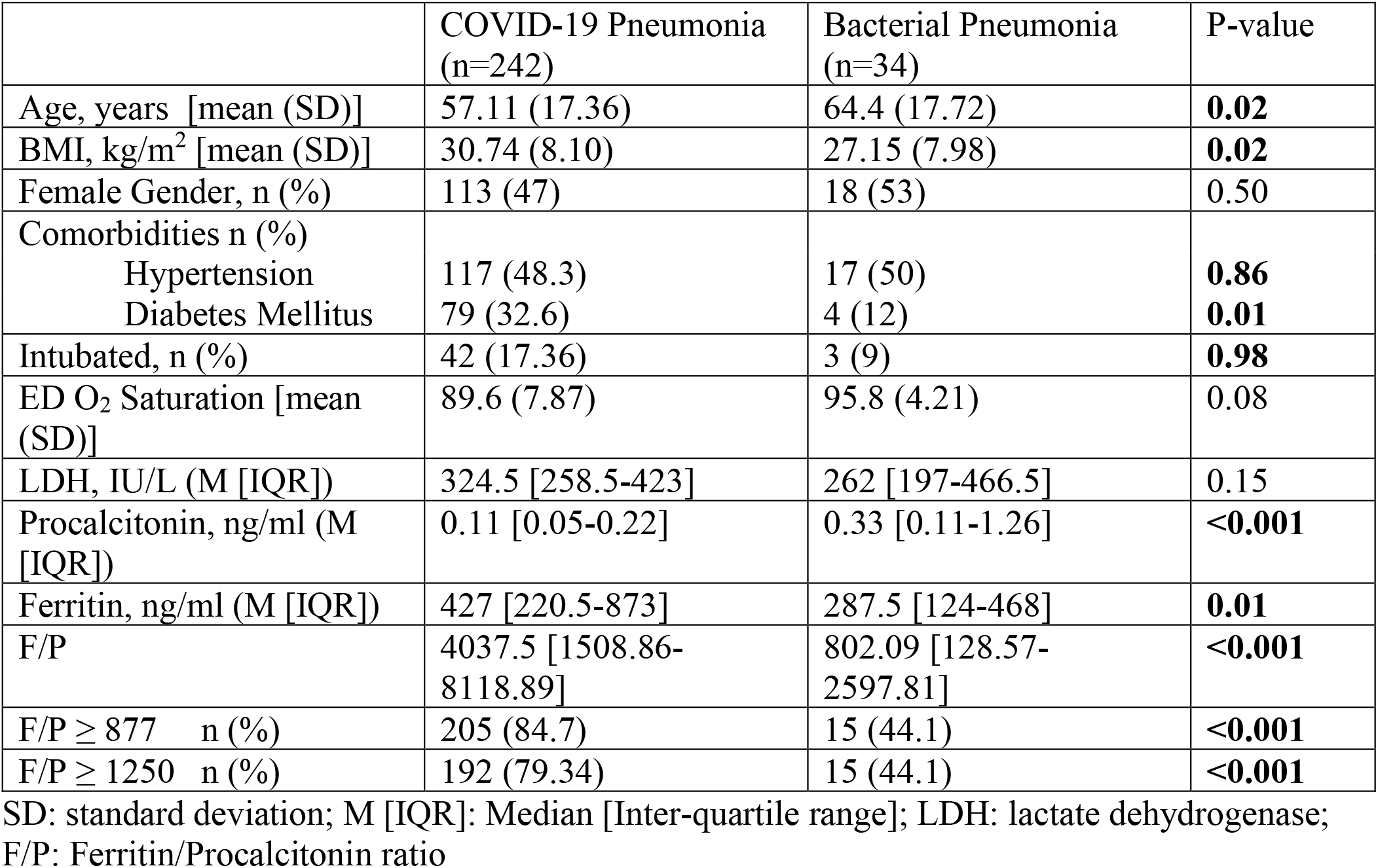
Characteristics of patients with COVID-19 pneumonia and bacterial pneumonia

### Ferritin, procalcitonin, and F/P

The median ferritin level was significantly higher in the COVID-19 pneumonia group compared to the bacterial pneumonia group. Patients with bacterial pneumonia had a significantly higher median procalcitonin level compared to patients with COVID-19 pneumonia. The median F/P was significantly higher in patients with COVID-19 (Table 2 and Figure 1B).

**Figure 1A:**
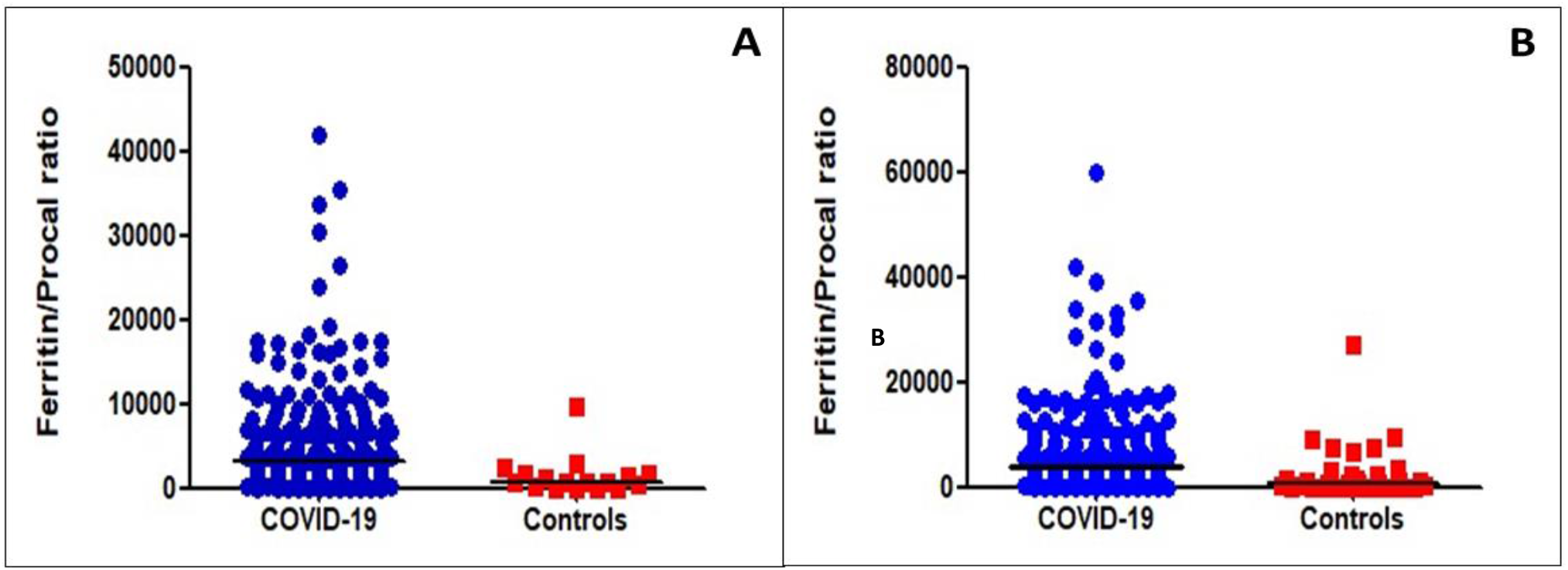
Ferritin to Procalcitonin Ratios of patients with COVID-19 pneumonia and patients with Bacterial Pneumonia (controls) at UCHealth. Figure 1B: Ferritin to Procalcitonin Ratios in patients with COVID-19 pneumonia and patients with Bacterial Pneumonia (controls) in the three cohorts combined.

### Predictors of COVID-19 Infection

Adjusting for age, gender, BMI, DM, HTN, and hospital, an F/P ≥ 877 was associated with an eleven-fold increased odds of COVID-19 compared to bacterial pneumonia (OR=11.27; CI [4.07-31.20]; p<0.001) (Table 3).

**Table 3:** Predictors of COVID-19 pneumonia in multivariate logistic regression

| Parameter | OR (95% CI) | P-value |
| --- | --- | --- |
| F/P ≥ 877 | 11.27 (4.07-31.20) | **<0.001** |
| Age | 0.98 (0.95-1.01) | 0.16 |
| Gender | 0.36 (0.14-0.98) | **0.05** |
| BMI | 1.06 (0.99-1.13) | 0.07 |
| DM Type 2 | 2.66 (0.78-9.04) | 0.12 |
| Hypertension | 0.47 (0.16-1.38) | 0.17 |
| Hospital | 0.45 (0.25-0.80) | **0.006** |

### Using the separate cohorts to assess the consistency of the F/P ratio

The F/P was assessed independently in the UCHealth patients and the combined UCHealth, Georgia, and China patients. A total of 218 patients with COVID-19 and 17 patients with bacterial pneumonia were diagnosed at UCHealth. The median F/P in the UCHealth group was significantly higher in patients with COVID-19 pneumonia than in the bacterial pneumonia group (3195 vs 860, respectively, p<0.001, Figure 1A). An F/P cut-off point of ≥ 1250 generated a sensitivity of 78% and a specificity of 59% to separate COVID-19 cases from bacterial pneumonia cases, with an AUC of 0.78 (Figure 2A). Adjusted for age, gender, BMI, DM, and HTN, an F/P ≥ 1250 associated with COVID-19 (OR: 4.9, CI: 1.5-16.1, p=0.009).

**Figure 2A:**
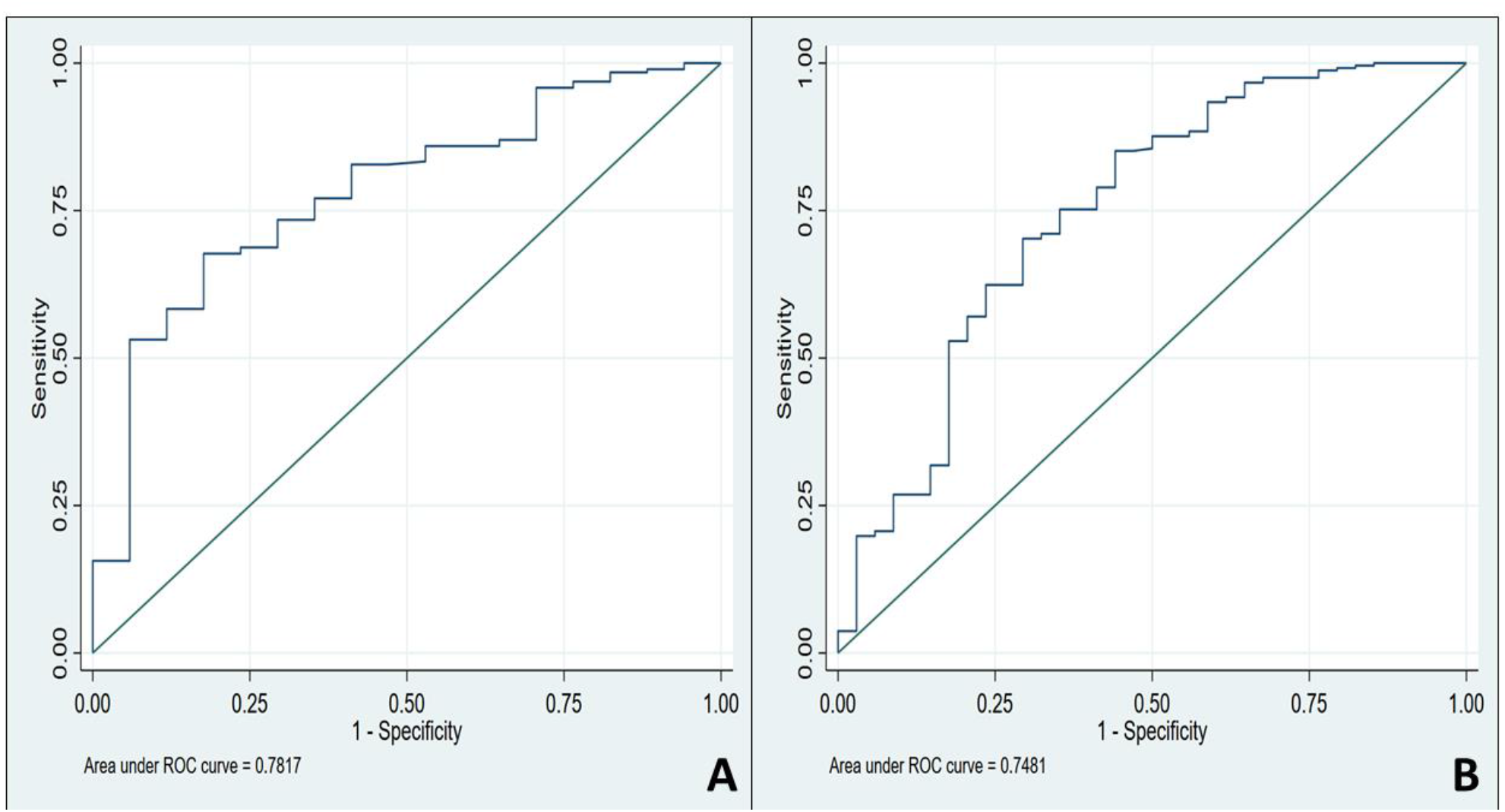
Receiver Operating Characteristic Analysis of Ferritin to Procalcitonin Ratio Cut-off Values Predicting COVID-19 Diagnosis for UCHealth. Figure 2B: Receiver Operating Characteristic Analysis of Ferritin to Procalcitonin Ratio Cut-off Values Predicting COVID-19 Diagnosis in the three cohorts combined.

Using data from the three participating sites combined, we found that a cut-off point ≥ 877 generated a sensitivity of 85% and a specificity of 56% to separate COVID-19 from bacterial infection, with an area under the curve (AUC) of 0.75 (Figure 2B). Adjusted for age, gender, BMI, DM, and HTN, an F/P ≥ 877 associates with a COVID-19 case (OR=11.27; CI [4.07- 31.20]; p<0.001) (Table 3). The positive predictive value of an F/P ≥ 877 was 93.2% for COVID-19 pneumonia.

## Discussion

In this retrospective multicenter study, an F/P cut-off point of ≥ 877 generates a sensitivity of 85% and a specificity of 56% to differentiate COVID-19 pneumonia from bacterial pneumonia. We assessed the F/P performance in patients from UCHealth and separately in all three hospitals combined. The results obtained in the UCHealth cohort were similar to the combined three-institution cohort, suggesting the diagnostic value of the F/P generalizes to other patient populations.

We created the F/P index using a model-based approach to biomarker development. A model is a simplified description of a phenomenon (disease) using tools derived from background knowledge or theories.^7,8^ Model components comprise objects (SARS-CoV-2, bacteria, cytokines, cells) and cause-effect interactions (effect of replication on cells, effects of pathogen on inflammation, and effect of host inflammatory molecules on biomarkers). In our model, SARS-CoV-2 produces disease by virion replication and obligate cytolysis in the respiratory epithelium that results in respiratory dysfunction.^9^ The magnitude of virus replication and cytolysis reflects disease severity. There is also an associated inhibition of systemic inflammation caused by-products of viral replication. In contrast, in bacterial pneumonia, most organisms do not require intracellular replication and cytolysis and thus produce less respiratory cell destruction and more systemic inflammation compared to SARS-CoV-2. This model focuses on pathogen replication and host inflammation during infection, and we sought biomarkers that best represented these two model components. We restricted candidate biomarkers to those widely available, rapidly obtainable, and inexpensive.

The first model component we assessed was the role of cytolysis in infection. Significant cytolysis caused by SARS-CoV-2 replication is suggested by *in vitro* observations in the closely related virus SARS-CoV, where prodigious viral replication within human primary ciliated respiratory epithelial cells caused cytopathy and cell shedding from tissues.^10^ Ferritin stores iron within cells and protects cell components from iron-induced free radical generation.^11^ Circulating ferritin likely originates from cell injury or cytolysis.^12^ Observations supporting this concept include the absence of a mechanism for cell secretion of ferritin.^12^ Since serum ferritin levels are unaffected by elevated plasma tumor necrosis factor-alpha (TNF) or Interleukin-6 (IL-6) in healthy volunteers, serum ferritin is unlikely to track inflammation and is probably not an acute phase reactant.^13^ A close correlation has been shown between serum ferritin levels and circulating cell-free DNA (cfDNA) in hemodialysis patients.^14^ Cell-free DNA in blood derives from cytolysis or cell apoptosis and is a cytolysis footprint that appears to be mirrored by elevated ferritin.^15^ Cell-free DNA is elevated in COVID-19, and increased cfDNA is associated with increased disease severity.^16^ This suggests a role for significant cytolysis in COVID-19 pathogenesis. These data strongly suggest ferritin in blood originates from cytolysis, and circulating ferritin levels reflect the magnitude of cytolysis. The association of viral infection of cells with cytolysis and liberation of intracellular ferritin into the blood is consistent with substantial elevation of blood ferritin reported in COVID-19 patients.^3,17,18^ Since the exact source of circulating cfDNA and ferritin during COVID-19 is unknown, lysis of non-respiratory epithelial cells may be additional sources for these cytolysis products. Regardless of cell source, the magnitude of total cytolysis in COVID-19 patients likely reflects total viral replication activity and indicates disease severity. In contrast to COVID-19, bacterial pathogenesis does not necessitate host cell lysis since most typical bacterial pneumonia pathogens do not require intracellular invasion to replicate. We believe it is useful to think of circulating ferritin as a biomarker indicating cytolysis and suggest an analogy with circulating levels of cardiac isoenzymes (often Troponin proteins) as indicators of cardiomyocyte lysis or damage.

Inflammation is the second major component of our model. TNF and IL-1β (IL-1) are prototype pro-inflammatory mediators involved in the host immune response. TNF levels are significantly lower in patients with COVID-19 and ARDS compared to patients with bacterial septic shock with ARDS,^19^ suggesting a less robust inflammatory response in COVID-19 compared to bacterial infections. Procalcitonin (PCT) is another marker of inflammation and has been shown to increase in response to TNF and IL-1.^20^ Moreover, PCT synthesis is suppressed by interferon-gamma (IFN), which is produced in response to viral infections and may participate in lowering PCT levels during viral infections.^20^ A review comparing PCT levels in viral and bacterial pneumonia in pediatric patients showed lower PCT concentrations in viral pneumonia.^21^ For adult pneumonia, a systematic review and meta-analysis revealed PCT >0.05 μg/mL used to diagnose bacterial infection is associated with sensitivity and specificity of 0.55 and 0.76 respectively (considered a moderate degree of discrimination).^22^ Considered together, it appears viral pneumonia —like COVID-19— produces lower systemic inflammation compared to bacterial pneumonia, and this is reflected in lower PCT concentrations in viral pneumonia. Given its widespread availability, lower cost, and fast turnaround time compared with levels of TNF or IL-1, we chose PCT as a surrogate marker of inflammation.

The F/P may have substantial clinical applicability. Circulating ferritin and procalcitonin levels can be easily obtained on routine blood testing upon admission, with results providing real-time information on whether patients are more likely to have COVID-19 compared to bacterial pneumonia. This information may guide antimicrobial administration, potential isolation until the diagnosis of COVID-19 is confirmed, and specific treatment considerations. Unnecessary antimicrobial therapy and the harms associated with its use can be spared. The prevalence of bacterial co-infections among patients with COVID-19 in our study was 15.4%, and we excluded these patients from this report. In a large cohort of 1705 COVID-19 patients, the rate of community-onset bacterial co-infections was only 3.5%,^23^ and 4,267 COVID-19 patients in New York City co-infections were diagnosed in 3.6% of the cases.^5^ The larger percentage of co-infected patients in our study in the total patient group is a reflection of patient population variability. Although not addressed in this report, we are evaluating F/P for use in discerning co-infected patients from patients infected with COVID-19 alone.

This study has several limitations. Proper use of F/P requires obtaining ferritin and PCT simultaneously as soon as possible after clinical presentation. However, our data included the first available levels of ferritin and PCT, and these levels may not have been obtained simultaneously or immediately after initial presentation. The diagnostic characteristics of F/P may improve with use of early and simultaneous acquisition of ferritin and PCT. Our total patient cohort from all 3 hospitals included only 34 patients with bacterial pneumonia, and more bacterial pneumonia cases need to be assessed to obtain a more accurate estimate of the utility of F/P. The retrospective study design makes it susceptible to several biases and the applicability of F/P to other patient populations is uncertain. Due to limitations of available data, we were unable to assess F/P association with important outcomes like mortality, ICU stay, and mechanical ventilation. Although we identified and adjusted for several confounders, there may be confounders unaccounted for in our analysis. Furthermore, we observed F/P overlap at low values for F/P (Fig 1), and our model predicts this is likely due to COVID-19 patients with mild disease. Low F/P in COVID-19 likely associates with smaller amounts of cytolysis and increased inflammation. It may therefore be difficult to separate mild COVID-19 cases with cases of bacterial pneumonia. If our model accurately reflects disease mechanism, serial calculation of F/P could be used as a tool to quantify the severity of infection over time and may predict clinical outcomes like mortality. Finally, we are uncertain how co-infection with SARS-CoV-2 and bacterial pathogens affects F/P.

In summary, the novel F/P may assist with early identification of COVID-19 patients, and further studies should evaluate its potential to quantify COVID-19 severity and prognosis.

## Data Availability

The datasets used in the current study are available from the corresponding author on a reasonable request.

## Acknowledgements

We acknowledge the support of the University of Colorado medical students who contributed significantly to the chart abstraction.

